# Enhancing Genetic Association Power in Endometriosis through Unsupervised Clustering of Clinical Subtypes Identified from Electronic Health Records

**DOI:** 10.1101/2024.04.22.24306092

**Authors:** Lindsay Guare, Leigh Ann Humphrey, Margaret Rush, Meredith Pollie, Yuan Luo, Chunhua Weng, Wei-Qi Wei, Leah Kottyan, Gail Jarvik, Noemie Elhadad, Penn Medicine Biobank, Regeneron Genetics Center, Krina Zondervan, Stacey Missmer, Marijana Vujkovic, Digna Velez-Edwards, Suneeta Senapati, Shefali Setia-Verma

## Abstract

**Background:** Endometriosis affects 10% of reproductive-age women, and yet, it goes undiagnosed for 3.6 years on average after symptoms onset. Despite large GWAS meta-analyses (N > 750,000), only a few dozen causal loci have been identified. We hypothesized that the challenges in identifying causal genes for endometriosis stem from heterogeneity across clinical and biological factors underlying endometriosis diagnosis.

**Methods:** We extracted known endometriosis risk factors, symptoms, and concomitant conditions from the Penn Medicine Biobank (PMBB) and performed unsupervised spectral clustering on 4,078 women with endometriosis. The 5 clusters were characterized by utilizing additional electronic health record (EHR) variables, such as endometriosis-related comorbidities and confirmed surgical phenotypes. From four EHR-linked genetic datasets, PMBB, eMERGE, AOU, and UKBB, we extracted lead variants and tag variants 39 known endometriosis loci for association testing. We meta-analyzed ancestry-stratified case/control tests for each locus and cluster in addition to a positive control (Total N_endometriosis cases_ = 10,108).

**Results:** We have designated the five subtype clusters as pain comorbidities, uterine disorders, pregnancy complications, cardiometabolic comorbidities, and EHR-asymptomatic based on enriched features from each group. One locus, *RNLS*, surpassed the genome-wide significant threshold in the positive control. Thirteen more loci reached a Bonferroni threshold of 1.3 x 10^-3^ (0.05 / 39) in the positive control. The cluster-stratified tests yielded more significant associations than the positive control for anywhere from 5 to 15 loci depending on the cluster. Bonferroni significant loci were identified for four out of five clusters, including *WNT4* and *GREB1* for the uterine disorders cluster, *RNLS* for the cardiometabolic cluster, *FSHB* for the pregnancy complications cluster, and *SYNE1* and *CDKN2B-AS1* for the EHR-asymptomatic cluster. This study enhances our understanding of the clinical presentation patterns of endometriosis subtypes, showcasing the innovative approach employed to investigate this complex disease.

## Introduction

Endometriosis, a complex gynecological condition affects 10% of women of reproductive age globally and more than 50% of women with infertility (1), yet it often goes either undiagnosed or misdiagnosed, leading to delayed diagnoses and delivery of effective therapy (2,3). Endometriosis is primarily characterized by the presence of endometrial-like tissue outside of the uterus. For managing the condition without surgery, the main treatments include pain relief and hormone-based therapies, neither of which are curative. A notable number of women with endometriosis receive opioids for pain management, despite the need for more sustainable and effective treatment options (4,5). On the other hand, hormonal therapies may have limitations to utilization due to severe side effects or a desire to become pregnant. Typically, the treatment for endometriosis often includes both medical and surgical approaches, however 30-50% of patients with severe endometriosis may require a second surgery within 3-5 years (6). The most comprehensive surgical management involves a hysterectomy with bilateral salpingoophorectomy (7).Treatment and health care visits accumulate many direct and indirect costs for women with endometriosis. The estimated economic cost of endometriosis in the US is ∼$10k per patient which is ∼14% higher than that of diabetes (8), and does not include the costs patients incur by having to miss work days because of their symptoms. In total, endometriosis presents a high economic burden that exceeds $22 billion in the U.S. alone (9). The condition not only imposes significant costs but also involves severe symptoms, delayed diagnosis, limited treatment options, and financial strain: challenges that could be significantly mitigated with a more detailed understanding of the disease.

Electronic Health Records (EHRs) represent a rich, yet underutilized, data source for capturing the phenotypic spectrum of endometriosis (10). Although the symptoms for endometriosis can be quite severe, including chronic debilitating pain, dyspareunia, and infertility, the average time to diagnosis is 4.5 years (11), in part because the only way to definitively diagnose endometriosis is by surgical observation of endometrial lesions growing outside of the uterus (e.g. abdominal cavity, pelvis, ovaries, etc.) (12). The variability in symptoms and disease presentation adds to the difficulty of diagnosis and hinders the optimal use of electronic health records (EHRs) in research for accurately identifying affected individuals and control subjects (13–15), which is critical for understanding the disease and advancing treatment strategies. The depth and breadth of EHR data provide a unique opportunity to apply unsupervised learning techniques for the identification of distinct phenotypic clusters that may correspond to clinical subtypes of endometriosis. Such an approach aligns with precision medicine’s goal to tailor diagnosis and treatment strategies to individual patient characteristics, potentially revealing novel insights into the disease’s pathophysiology.

Better understanding of the disease mechanisms of endometriosis could lead to improved diagnostic practices, reducing costs to the healthcare system and improving quality of life through treatment and earlier diagnosis for patients. In spite of the prevalence and severity of endometriosis, etiology of endometriosis is still poorly understood. The pursuit thus far of biomarkers and drug targets based on genetic contributions of disease in patients with endometriosis has mainly included genome-wide association studies to identify genetic variants contributing to the disease (16,17). Twin studies have estimated the heritability of endometriosis to be 47.5% (18), and common variants are estimated to contributed 26% of phenotypic variance (19), but the largest GWAS to-date (N > 750,000, 60,674 cases) has only explained 9% of the phenotypic variance (17). Although these recent advances in genomic studies have promised insights into the underlying genetic mechanisms of endometriosis, yet the heterogeneity of the disease presentation has consistently complicated these efforts. Traditional genetic association studies have struggled to untangle the intricate web of genotypic and phenotypic diversity within endometriosis patients, leading to a critical need for innovative approaches to dissect the disease’s complexity.

We hypothesized that underlying clinical heterogeneity is obscuring the genetic mechanisms and preventing large-scale genetic studies from explaining more of the heritability. Endometriosis causes a wide range of symptoms and concomitant conditions, including severe chronic pain, gastrointestinal inflammation, and infertility. Additionally, many symptoms of endometriosis are shared between other gynecological diseases such as primary dysmenorrhea, ovarian cysts, and pelvic inflammatory disease; making symptom-based diagnosis challenging (20,21). Recent studies have highlighted the importance of complex disease subtyping in improving our understanding of the genetic mechanisms underlying endometriosis. For example, a recent study on polycystic ovary syndrome (PCOS) used unsupervised clustering to identify three subtypes of PCOS based on lab and biometric values before conducting genome-wide association study for each subtype (22). This approach allowed for a more nuanced understanding of the genetic basis of PCOS and could be applied to endometriosis to identify subtypes with distinct genetic mechanisms. Building on the premise that a more nuanced understanding of endometriosis subtypes could unlock new genetic associations, our study leverages unsupervised, phenotypic clustering analysis of EHR data to systematically identify and characterize clinical subtypes of endometriosis. By dissecting the heterogeneity inherent in the disease, we aim to increase the power of genetic association analyses, facilitating the identification of subtype-specific disease mechanisms. This approach not only promises to enhance our understanding of endometriosis genetics but also to refine diagnostic criteria and inform more targeted and effective treatment strategies.

In conclusion, the complex nature of endometriosis, with its diverse symptoms and overlapping features with other gynecological diseases, presents challenges for understanding its genetic mechanisms. In this manuscript, we detail the methodology and findings of our study, which integrates unsupervised phenotypic clustering with subsequent genetic association analyses for each identified endometriosis subtype. By doing so, we aim to bridge the gap between clinical observations and genetic research in endometriosis, providing a roadmap for future studies to explore the genetic underpinnings of this complex disease with renewed clarity and precision. This deeper understanding may pave the way for more targeted and personalized approaches to diagnosis, treatment, and management of this debilitating condition. Further research and large-scale genetic studies are needed to fully elucidate the genetic architecture of endometriosis and its subtypes, ultimately leading to improved outcomes for affected individuals.

## Methods

### Datasets Used for Sub-phenotyping and Genetic Association

The Penn Medicine Biobank (PMBB) is the University of Pennsylvania’s health system-based biobank which consists of about 250,000 consented participants, with 43,624 of those having imputed genotype data (imputed to TOPMED reference panel) linked with their electronic health record (EHR) history. The PMBB is an electronic health record (EHR)-linked biobank that integrates a wide variety of health-related information, including diagnosis codes, laboratory measurements, imaging data, and lifestyle information, with genomic and biomarker data. The PMBB is one of the most diverse medical biobanks, with approximately 30% of participants being of non-European ancestry. This diversity is crucial for ensuring that research findings are applicable to a broad range of populations. The biobank also benefits from a median of seven years of longitudinal data in the EHR, providing valuable information on participants’ health histories (22). For our study, we treated the PMBB as two distinct datasets: those without and those with genotype data. EHR data from the non-genotyped PMBB were used for cluster derivation whereas the genotyped PMBB cohort was used in the genetic analyses.

The Electronic Medical Records and Genomics (eMERGE) network is a National Human Genome Research Institute-funded consortium engaged in the development of methods and best practices for using the electronic medical record as a tool for genomic research. The eMERGE network is a publicly-available dataset with contributions from multiple health systems within the United States which contains about 100,000 participants with linked health records and imputed genomic data (imputed to HRC reference panel) (23). The eMERGE consortium validated the hypothesis that clinical data derived from electronic medical records can be used successfully for complex genomic analysis of disease susceptibility across diverse patient populations (24). The eMERGE network has shown the efficiency that can result from the use of electronic health record data.

The All of Us (AOU) Research Program is an initiative created by the NIH to recruit demographically diverse individuals to the largest US-based biobank to-date. Recruitment began in 2018, and since then, over 400,000 people have signed up and submitted baseline questions (25). 245,388 of them have short-read whole genome sequence data, collectively representing over one billion genetic variants (26). Participants’ EHRs are contributed to the AOU data processing center using the Sync for Science platform (27), which works with EHR vendors such as Epic and Cerner to collate structured patient data for research use (28).

The UK Biobank (UKBB) is a large and comprehensive dataset that provides valuable resources for researchers studying a wide range of health-related topics. The UKBB is a population-based publicly available dataset consisting of about 500,000 UK citizens with EHR data, health survey data, and imputed genotypes. The UK Biobank has performed genome-wide genotyping on all participants using the UK Biobank Axiom Array (29). This array directly measures approximately 850,000 variants, and more than 90 million variants are imputed using the Haplotype Reference Consortium and UK10K + 1000 Genomes reference panels.

All four of the biobanks mentioned above (PMBB, eMERGE, AOU, and UKBB) utilize the Observational Medical Outcomes Partnership Common Data Model (OMOP-CDM) to represent structured EHR data in a harmonized format (30). For this study, we utilized women with ICD-diagnosed endometriosis in the non-genotyped PMBB cohort (N_endo_ = 4,078) as the derivation dataset for the clinical subtypes. For deeper characterization of our subtypes, we performed chart-reviews on 682 randomly selected endometriosis cases from the genotyped PMBB. Then, we meta-analyzed women from the genotyped PMBB (N = 20,697, N_endo_ = 1,198), six non-pediatric sites within the eMERGE network (N = 51,800, N_endo_ = 2,243), the AOU research program (N = 108,098, N_endo_ = 2,126), and UKBB (N = 261,824, N_endo_ = 4,451) to form our main genetic analysis test set (N = 442,419, N_endo_ = 10,018).

Each of the biobanks projected their samples onto the thousand genomes reference population and performed clustering to assign genetically inferred ancestry labels corresponding to those from the thousand genomes project (31). We restricted our genetic association analyses to the groups which had substantial sample sizes, which were those with high similarity the AFR and EUR thousand genomes superpopulations. We will refer to those groups using AFR and EUR from here on out.

### Extraction of Endometriosis-Related Clinical Features

Patients with endometriosis have heterogeneous clinical presentations; there are a wide variety of associated symptoms, risk factors, and comorbidities. We first determined participants’ case-control status of endometriosis using structured EHR data: ICD-9 and ICD-10 billing codes 617 and N80, respectively. Then for endometriosis cases, we determined whether each individual had a history of endometriosis-related clinical features. In total, we extracted 39 ICD-based features (Table S1): 9 ICD-based anatomical subtypes, 14 comorbidities, 8 symptoms, and 8 pregnancy-related phenotypes. We selected only symptoms, comorbidities, and pregnancy-related conditions for clustering, removing the 9 anatomical subtypes to be used downstream in cluster characterization. We further restricted these conditions to those with a prevalence amongst endometriosis cases in the subtype dataset of at least 5%, leaving us with 17 features for the clustering analysis (Figure S1).

### Unsupervised Clustering

We tested four popular methods for unsupervised clustering: spectral clustering, density-based spatial clustering of applications with noise (DBSCAN), hierarchical agglomerative clustering, and k-means clustering. Spectral clustering identifies clusters by decomposing a dataset’s affinity matrix into its eigenvectors and then clustering in the eigenvector space using QR clustering algorithm (32,33). DBSCAN is an algorithm which identifies dense regions of data points to discover clusters (34). Hierarchical agglomerative clustering is an unsupervised classification method that uses a pairwise distance matrix to iteratively merge nearby points together (35). K-means clustering randomly initializes centroids for each cluster and then alternates between assigning data points to their nearest centroid and adjusting the centroids until convergence (36).

In addition to choosing an algorithm, a common struggle with unsupervised clustering is choosing a target number of clusters in a non-arbitrary way. We used several empirical metrics for this: silhouette score, distortion score, and a metric we developed to represent the “evenness” of clusters. The silhouette score is a metric which considers both intra- and inter-cluster distances to assess tightness within a cluster and distance between clusters; higher silhouette scores indicate better quality clusters. The distortion score is the sum of squared errors with respect to the centroid of each cluster, thus it is desired to minimize distortion. Our evenness metric, optimized by minimization, was defined as the fractional difference between the size of the largest and smallest clusters. We measured these metrics across tests for 2-20 clusters for each of the four clustering methods (except for DBSCAN which automatically infers the optimal number of clusters).

### Characterization of Unsupervised Clusters

After identifying clinical clusters within our observation dataset, our objective was to delineate their characteristics. We performed two-population z-score proportion tests (37) to determine if the rates of input conditions were significantly different on a cluster-vs-other-clusters basis. For our training set (the non-genotyped PMBB, N_endo_ = 4,078), we examined two sets of features for the z-score tests: the 17 input features as well as ICD-based anatomical subtypes of endometriosis including adenomyosis, endometrioma, superficial lesions, and deep lesions (Supplementary Table S2). For characterizing our clusters, we also utilized a chart-reviewed dataset of 682 genotyped PMBB patients with endometriosis ICD codes. The features considered here were confirmed endometriosis and adenomyosis status and chart-abstracted symptoms, comorbidities, and surgical phenotypes (Supplementary Table S3). By considering the cluster-specific differences in these EHR-derived features among the two datasets, we could observe patterns in clinical presentation. Based on these patterns, we assigned labels to each cluster.

### Cluster-Stratified Candidate Gene Association Testing

To identify genetic heterogeneity among the varied clinical presentations of endometriosis, we performed cluster-stratified, ancestry-stratified candidate gene association studies. Using PLINK 2.0 (38), we extracted single nucleotide polymorphisms (SNPs) in LD (kb distance < 0.5 Mb and R^2^ > 0.1) with 39 autosomal lead SNPs reported in the most recent endometriosis GWAS(17). LD was computed based on the thousand genomes reference panel (31). Cluster phenotypes were assigned for PMBB, eMERGE, and AOU using a K-Nearest neighbors’ classifier (39) with K=3 on the same 17 ICD-based features. For each study, we employed a linear mixed model regression method employed in SAIGE (40) to test for associations between genotypes and case-control status. Cases were females with endometriosis from one cluster and controls were biological females with no ICD history of endometriosis. In the regression models we included the first four principal components, age, and batch indicators (eMERGE only) as covariates. The ancestry-stratified results of these studies were then meta-analyzed using Plink 1.9 (41) for each of the cluster-phenotypes. We also tested a baseline overall endometriosis (cases from all clusters combined) as a positive control to identify how many known loci we were able to replicate. Because multiple genetic ancestry groups were included, we chose a random-effects meta-analysis, which is more robust to heterogeneity (42).

## Results

### Derivation, Study, and Validation Datasets

This study utilized five datasets to investigate the genetic mechanisms underlying endometriosis and its subtypes. The datasets used were endometriosis cases in the non-genotyped PMBB for the derivation of clusters, a chart-reviewed endometriosis cohort to help characterize the clusters, the genotyped PMBB, six sites within the eMERGE network, AOU, and UKBB for genome-wide association analyses (See Methods). The sample sizes for each cohort, the mean age at diagnosis, the number of cases and controls, and the mean age at the time of data pull for each cohort are shown in Table 1. See Methods for details on each of the four datasets. By leveraging these datasets, the study aimed to identify endometriosis subtypes and gain insights into the genetic factors associated with endometriosis and its subtypes.

**Table 1:** Cohort sample size and average age of cases and controls for the datasets used in this analysis. Age was considered the age as of when the EHR data were collected.

| Dataset | Endometriosis | N (AFR / EUR) | Mean Age (SD) |
| --- | --- | --- | --- |
| Cluster Derivation Set: |  |  |  |
| Non-Genotyped PMBB | Cases | 4,078 (NA) | 49.9 (13.3) |
| Genetic Association Sets: |  |  |  |
| AOU | Cases | 2,126 (542 / 1,584) | 52.2 (12.8) |
|  | Controls | 108,099 (31,435 / 76,664) | 56.8 (16.8) |
| eMERGE | Cases | 2,243 (353 / 1,890) | 59.9 (14.6) |
|  | Controls | 49,557 (9,934 / 39,623) | 59.7 (23.4) |
| PMBB | Cases | 1,198 (562 / 636) | 54.2 (12.9) |
|  | Controls | 19,493 (6,524 / 12,969) | 60.0 (17.8) |
| UKBB | Cases | 4,541 (112 / 4,429) | 51.5 (7.5) |
|  | Controls | 257,283 (4,524 / 252,759) | 56.6 (8.0) |
| Meta-Analysis Totals: |  |  |  |
| META | Cases | 10,108 (1,569 / 8,539) | 53.9 (11.3) |
|  | Controls | 434,432 (52,417 / 382,015) | 57.1 (13.6) |

### Derivation of Unsupervised of Clusters

Unsupervised clustering was performed in non-genotyped PMBB dataset of 4,078 women with EHR-diagnosed endometriosis using 17 clinical features (supplementary figure S2). We tested four methods for unsupervised clustering as well as 19 values for the number of clusters (K=2-20) and measured three metrics to empirically choose a clustering method and number of clusters (Figure 1).

**Figure 1:**
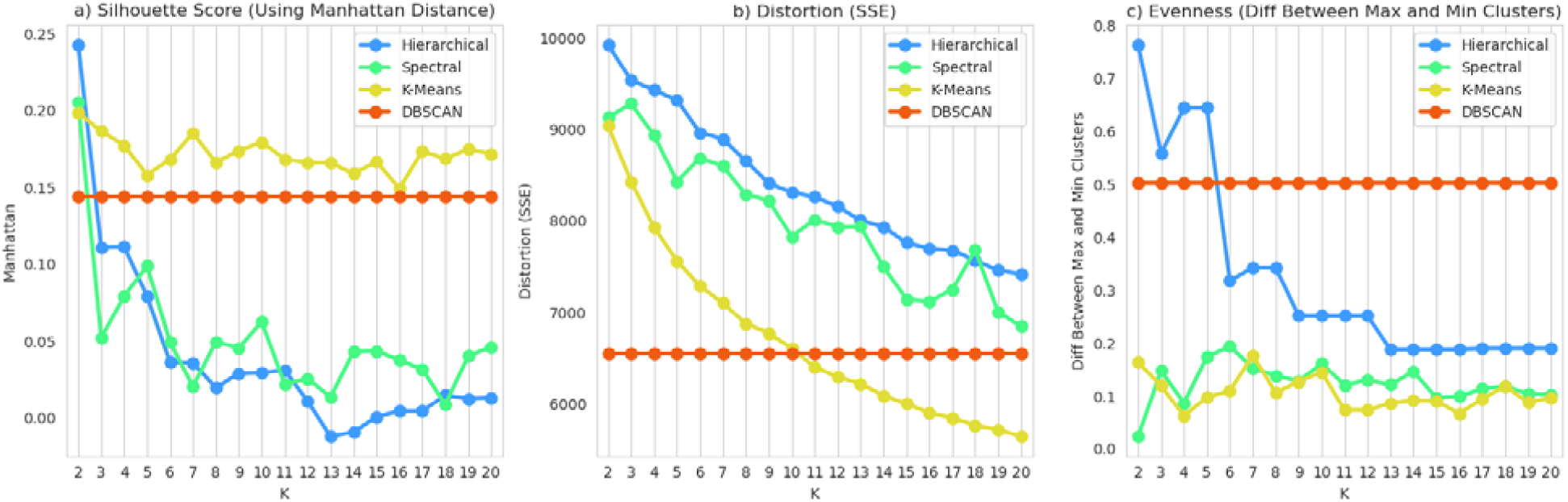
testing various clustering algorithms and K-values to empirically choose an optimal method. The three metrics shown are (a) Manhattan-distance-based silhouette score, (b) distortion or sum of squared errors, and (c) evenness represented by the difference in fraction between the largest and smallest clusters. Based on these tests, we chose spectral clustering with K=5.

Based on these tests, we first eliminated DBSCAN because the inferred number of clusters was 131, a far too complex model to be useful or interpretable. Next, we eliminated hierarchical clustering because the sizes of the resulting clusters were more uneven than the other methods. Spectral clustering and k-means clustering were ultimately more difficult to choose between, but when we focused on the shapes of the distortion curves across the values of K, we observed that k-means lacked an “elbow” to show a clear optimal K value whereas spectral clustering clearly indicated 5 as an ideal K with a local minimum. Thus, we chose spectral clustering with K=5 as our unsupervised subtyping model. The sizes of the final clusters were: (1) 441 - 11%, (2) 686 - 17%, (3) 1,151- 28%, (4) 796 - 20%, and (5) 1,004 - 25%. Figure 2 illustrates the eigenvectors of the affinity matrix which were used for clustering the data points.

**Figure 2:**
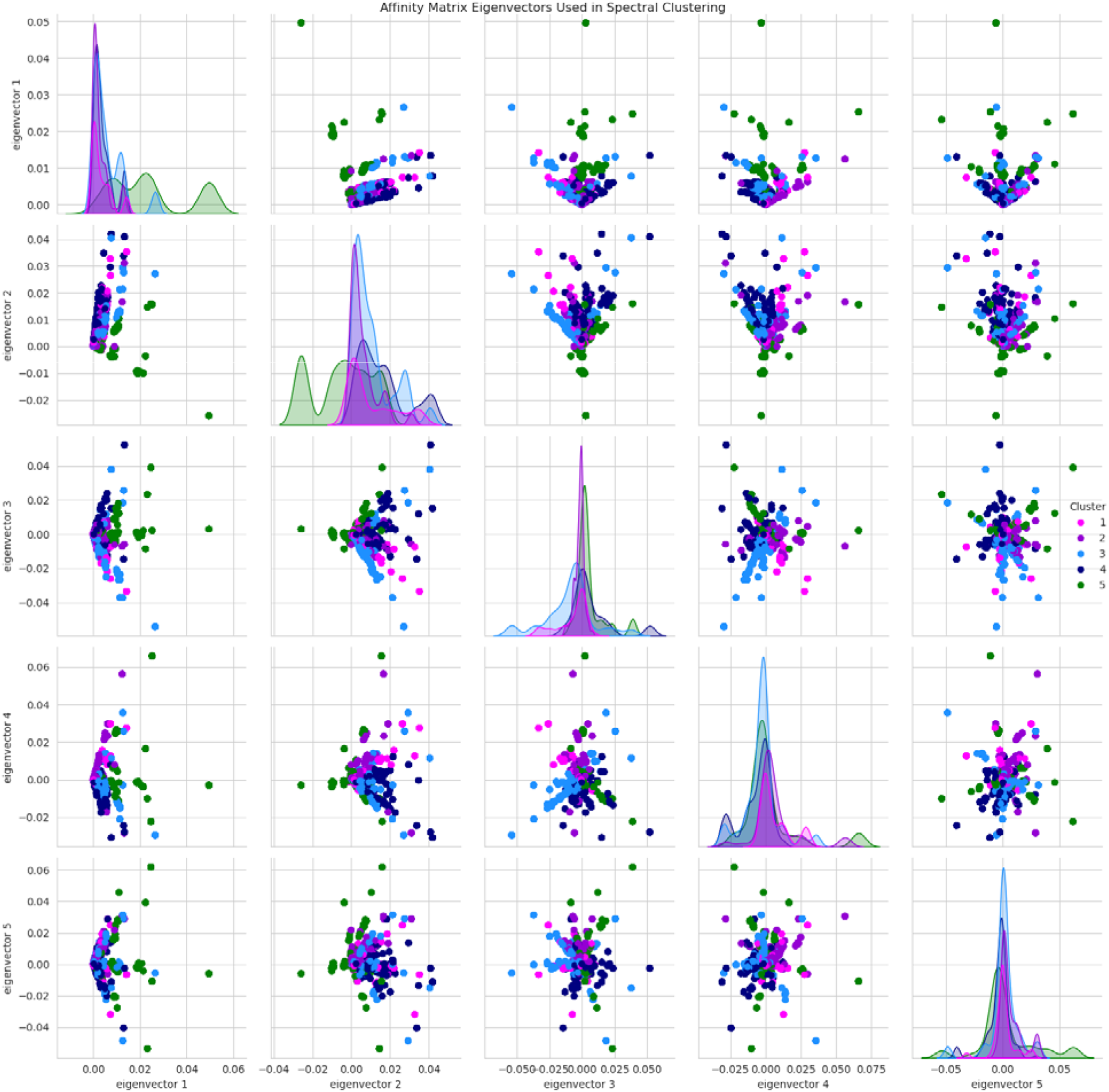
pairwise scatter plots of the first five eigenvectors of the affinity matrix used for spectral clustering, colored by cluster. This five-dimensional eigenvector space was used for clustering. The diagonal shows kernel density estimator plots for each of the five eigenvectors.

### Data-Driven Cluster Characterization

After clustering, we aimed to characterize these clusters by observing patterns in clinical presentation (prevalence) amongst the input features. We performed two sets of z-score proportion tests comparing prevalence of each feature between each cluster and the other four clusters in our training set. The first set of tests was performed on the original cluster derivation cohort, and the features included were the 17 input features (symptoms and comorbidities with prevalence > 5%) as well as ICD-defined anatomical subtypes of endometriosis (Figure 3).

**Figure 3:**
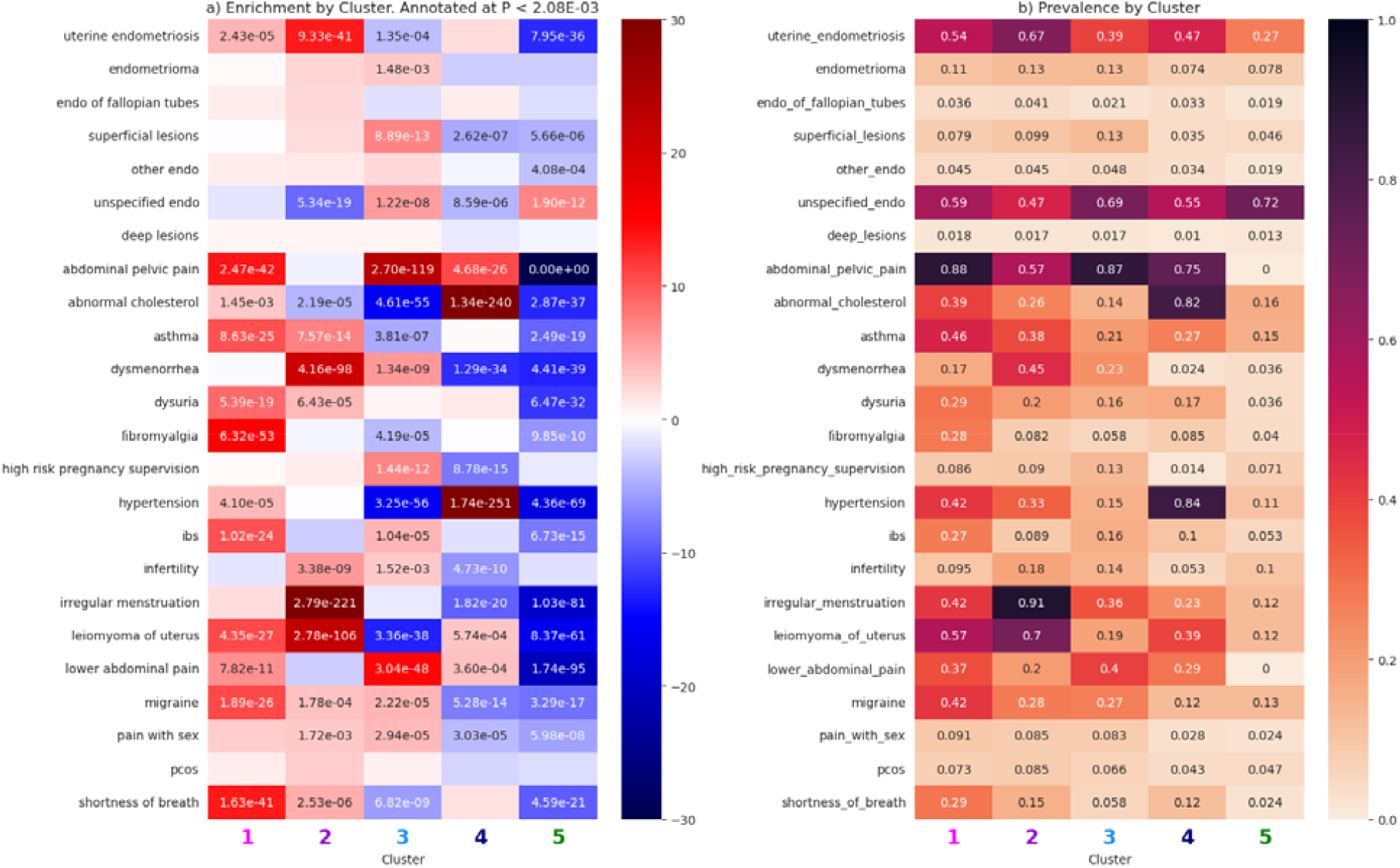
feature tests for the non-genotyped PMBB training set. Shown are (a) z-scores for the difference in proportion tests, annotated with p-values that are significant and (b) feature prevalence by cluster to provide context for the z-score tests.

Among the five clusters identified in the training set, there were many input features and ICD-based anatomical subtypes with significantly different proportions. To identify distinguishing features between the clusters, we focus on phenotypes which were significantly enriched and had the highest prevalence in that cluster. Cluster one had the highest rates of (and was significantly enriched for) dysuria (Z=8.9), migraine (Z=10.6), IBS (Z=10.3), fibromyalgia (Z=15.3), asthma (10.3), abdominal pelvic pain (Z=13.6), and shortness of breath (Z=13.5). Cluster two had the highest rates of the following significantly enriched traits: dysmenorrhea (Z=21.9), infertility (Z=5.9), irregular menstruation (Z=31.75), leiomyoma of uterus (Z=21.9), and uterine endometriosis defined by ICD-9 617.0* or ICD-10 N80.0* (Z=13.4). Cluster three’s defining features were high risk pregnancy supervision (Z=7.1), superficial lesions defined by ICD-9 617.3* or ICD-10 N80.3* (Z=7.1), and lower abdominal pain (Z=14.6). Individuals in cluster four had highest prevalence of abnormal cholesterol (Z=33.1) and hypertension (Z=33.9), while cluster five was only enriched for unspecified endometriosis defined as ICD-9 617.9* or ICD-10 N80.9* (Z=7.0).

The second set of tests was performed on a subset of endometriosis cases (N=682) from the genotyped PMBB for whom chart reviews were performed by OB-GYN clinical fellows at the University of Pennsylvania Hospital System. The features tested were gold standard confirmed diagnoses (endometriosis, adenomyosis, fibroids, and any ICD false positives), surgical subtypes, hormone use at the time of confirmation procedure, and symptoms identified from a combination of structured data and notes (Figure 4).

**Figure 4:**
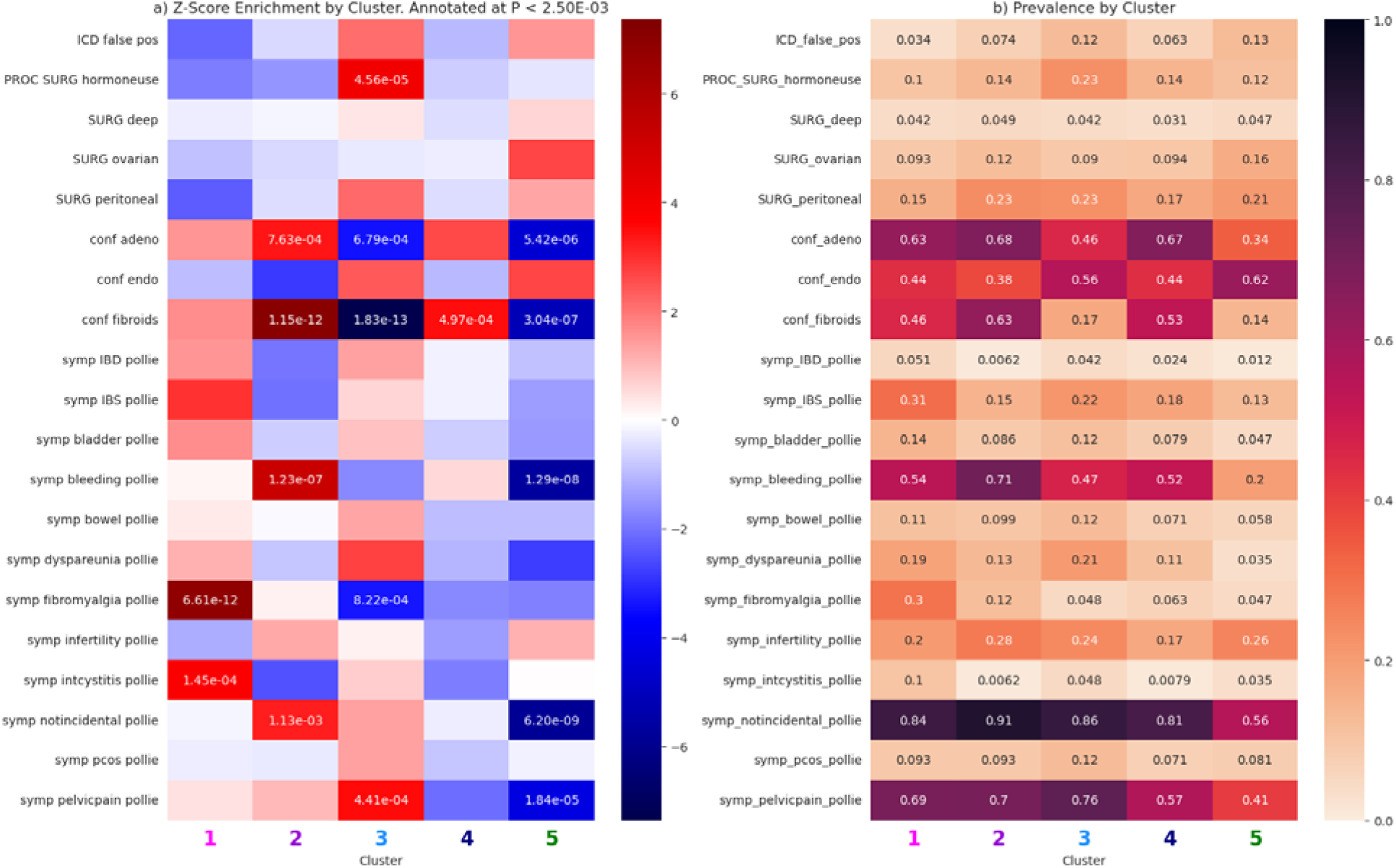
feature tests for the chart reviewed PMBB dataset. Shown are (a) z-scores for the difference in proportion tests, annotated with p-values that are significant and (b) feature prevalence by cluster to provide context for the z-score tests.

Because the size of our chart-reviewed dataset was limited, there were fewer significant tests. For cluster one, the phenotypes which were most significantly prevalent were interstitial cystitis (Z=3.8) and fibromyalgia (Z=6.9). For cluster two, the defining features were confirmed adenomyosis status (Z=3.7), confirmed uterine fibroids (Z=7.1), and symptomatic bleeding (Z=5.3). Cluster three’s most highly enriched features were pelvic pain (Z=3.5) and hormone use at the time of surgery (Z=4.1). Considering the enriched features for each cluster among the two sets of tests, we defined the following labels for 5 clusters: (1) pain comorbidities, (2) uterine disorders, (3 pregnancy complications, (4) cardiometabolic comorbidities, and (5) EHR-asymptomatic.

### Candidate Gene Association Testing Stratified by Phenotypic Cluster

We applied the subtype classifications observed in our derivation set to our four genetic association datasets, PMBB, eMERGE, AOU, and UKBB. We used a K-nearest neighbors model with the same 17 EHR-derived features to assign endometriosis cases to the five phenotypes (Table 2).

**Table 2:**
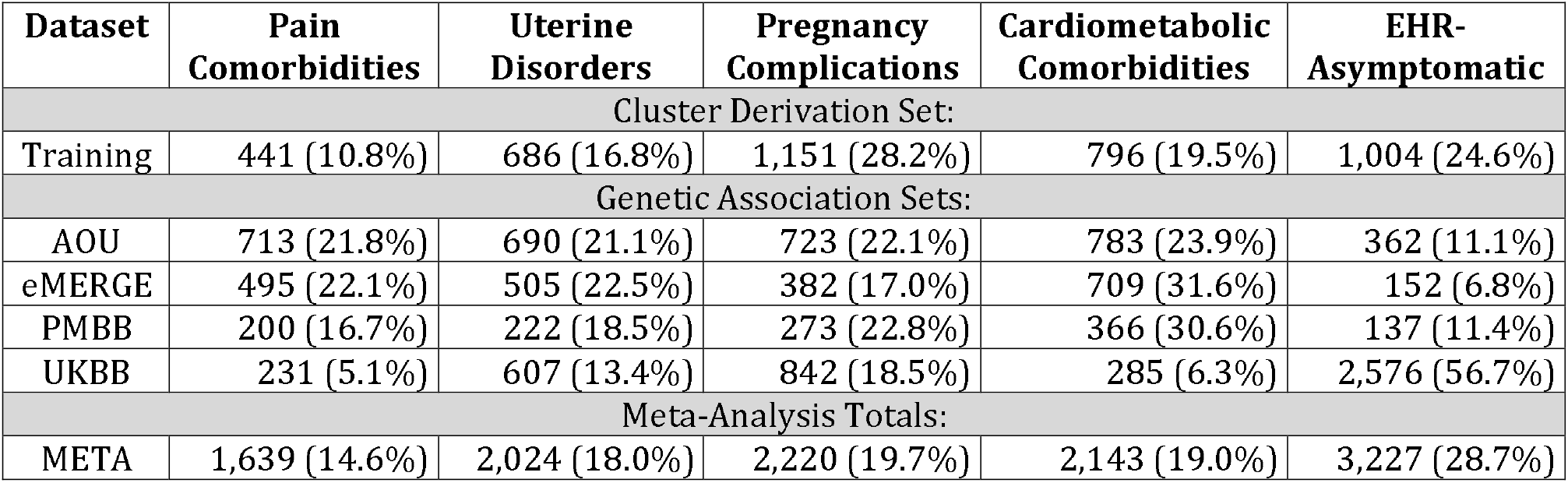
counts and proportions of endometriosis cases in each cluster by dataset.

| Dataset | Pain Comorbidities | Uterine Disorders | Pregnancy Complications | Cardiometabolic Comorbidities | EHR-Asymptomatic |
| --- | --- | --- | --- | --- | --- |
| Cluster Derivation Set: |  |  |  |  |  |
| Training | 441 (10.8%) | 686 (16.8%) | 1,151 (28.2%) | 796 (19.5%) | 1,004 (24.6%) |
| Genetic Association Sets: |  |  |  |  |  |
| AOU | 713 (21.8%) | 690 (21.1%) | 723 (22.1%) | 783 (23.9%) | 362 (11.1%) |
| eMERGE | 495 (22.1%) | 505 (22.5%) | 382 (17.0%) | 709 (31.6%) | 152 (6.8%) |
| PMBB | 200 (16.7%) | 222 (18.5%) | 273 (22.8%) | 366 (30.6%) | 137 (11.4%) |
| UKBB | 231 (5.1%) | 607 (13.4%) | 842 (18.5%) | 285 (6.3%) | 2,576 (56.7%) |
| Meta-Analysis Totals: |  |  |  |  |  |
| META | 1,639 (14.6%) | 2,024 (18.0%) | 2,220 (19.7%) | 2,143 (19.0%) | 3,227 (28.7%) |

The smallest cluster was the pain comorbidities cluster, with only 14.6% of total endometriosis cases being assigned to this cluster. The EHR-asymptomatic cluster was the largest cluster overall. The other three clusters occurred in relatively even proportions in the overall meta-analysis group at 18.0% (uterine disorders), 19.7% (pregnancy complications), and 19.0% (cardiometabolic comorbidities).

To establish a reference for the expected level of signal replication, we began with a positive control test. We conducted association tests on 39 established genetic locations (autosomes only) known to be linked to endometriosis. (Figure 5).

**Figure 5:**
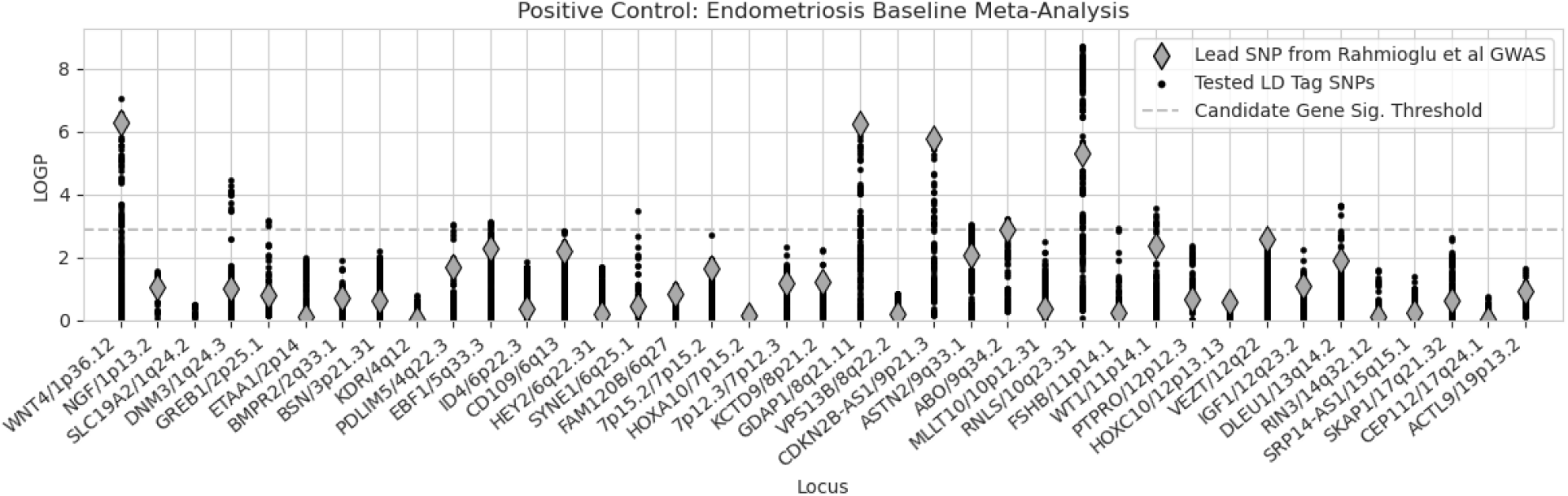
results for our endometriosis case vs control positive control association tests at each of the 39 known loci. Shown are the lead SNPs from the Rahmioglu et al 2023 GWAS as well their tag SNPs in LD (kb distance < 0.5 Mb and R^2^ > 0.1). X-axis labels are from the known GWAS.

Our positive control test resulted in fourteen replicating loci. Only one was genome-wide significant, *RNLS/10q23*.*31* (P = 1.91x10^-9^, rs792212:T). Thirteen were significant at a Bon Ferroni-corrected threshold of 0.05 / 39: *WNT4*/*1p36*.*12* (P = 9.12x10^-8^, rs2235529:T), *DNM3*/*1q24*.*3* (P = 3.54x10^-5^, rs655853:C), *GREB1/2p25*.*1* (P = 6.55x10^-4^, rs34532804:A), *PDLIM5*/*4q22*.*3* (P = 8.58x10^-4^, rs1493112:T), *EBF1/5q33*.*3* (P = 7.64x10^-4^, rs1878936:C), *SYNE1*/*6q25*.*1* (P = 3.20x10^-4^, rs13206045:C), *GDAP1*/*8q21*.*11* (P = 4.19x10^-7^, rs10957712:T), *CDKN2B-AS1*/*9p21*.*3* (P = 1.75x10^-6^, rs10122243:T), *ASTN2*/*9q33*.*1* (P = 9.01x10^-4^, rs62576127:A), *ABO*/*9q34*.*2* (P = 5.87x10^-4^, rs495828:G), *FSHB*/*11p14*.*1* (P = 1.17x10^-3^, rs11031006:A), *WT1*/*11p14*.*1* (P = 2.85x10^-4^, rs72638188:T), *DLEU1*/*13q14*.*2* (P = 2.24x10^-4^, rs9568417:G).

To test whether stratifying by clinical presentation allowed for greater resolution in genetic associations, we performed case-control candidate gene association studies for the five phenotypic clusters by meta-analyzing ancestry-stratified summary statistics from four EHR-linked genetic datasets: PMBB, eMERGE, AOU, and UKBB. We observe 18 / 39 loci (46%) significantly associating with one or more clusters (Figure 6a, 6b). Also, for up to 15 loci, the cluster-stratified phenotypes yield stronger associations than the positive control despite having smaller sample sizes (Figure 6c).

**Figure 6:**
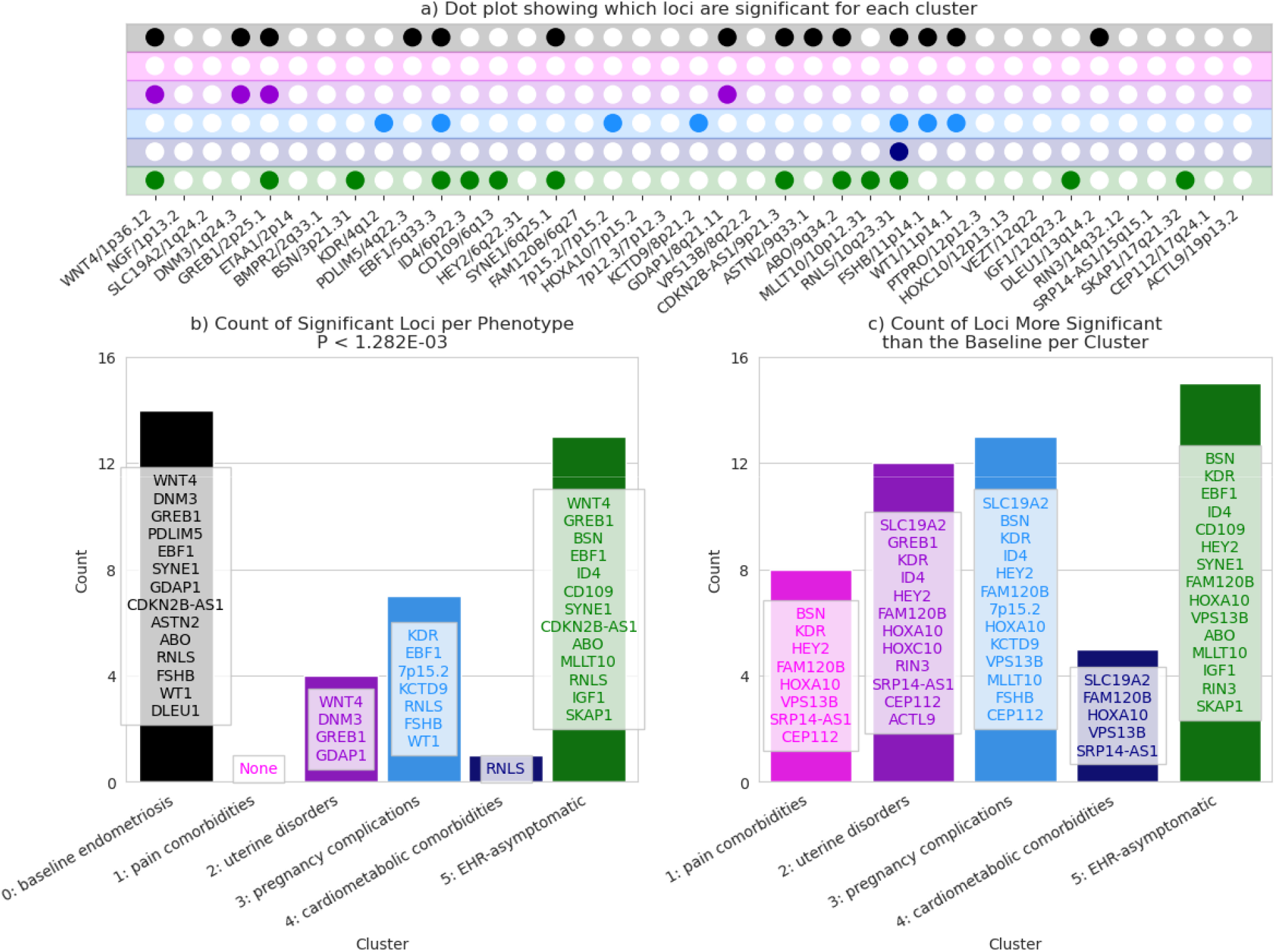
Phenotype-specific association test results. The top panel (a) indicates which known loci were significantly replicated by the positive control and five clusters. The bottom left panel (b) shows the number and names of statistically significant associations for each phenotype. The bottom right panel (c) shows the number of loci for which each phenotype had a more significant association than the baseline.

The smallest cluster, cluster one, with high rates of pain comorbidities, was not significantly associated with any known loci, but it was more significantly associated than the positive control for eight loci as shown in Figure 6c. The uterine disorders cluster (two) was significantly associated with four loci, *WNT4*/1p36.12, *DNM3*/*1q24*.*3, GREB1*/*2p25*.*1*, and *GDAP1*/*8q21*.*11*. Out of the seven loci significantly associated with the pregnancy complications cluster (three), three of them were not significantly associated with any other clusters or the positive control: *KDR/4q12, 7p15*.*2*, and *KCTD9/8p21*.*2*. Cluster four, enriched for cardiometabolic comorbidities, was significantly associated with one locus, *RNLS/10q23*.*31*, the strongest hit from the positive control. *RNLS* was also significantly associated with clusters three and five. Eleven loci were significantly associated with the EHR-asymptomatic cluster, and six of those (*BSN/3p21*.*31, ID4/6p22*.*3, CD109/6q13, MLLT10/10p12*.*31, IGF1/12q23*.*2*, and *SKAP1/17q21*.*32*) had no other associations, even with the positive control.

## Discussion

Endometriosis presents with heterogeneous symptoms ranging from severe pain to infertility, contributing to varying patient experiences and treatment responses. Several large genome-wide association studies and meta-analyses have been performed for endometriosis to-date. However, the genomic underpinnings of endometriosis remain incompletely understood, largely due to the clinical heterogeneity and the limitations of traditional genome-wide association studies (GWAS) that aggregate all cases into a single analysis pool. This approach may obscure genetic variations specific to different endometriosis phenotypes, thus necessitating more refined stratification techniques. There are various approaches to phenotyping participants for these studies including surgical notes and electronic health records. While there have also been analyses which account for disease progression (17), there have not been any genome-wide investigations into the genetics underlying the heterogeneous presentation patterns of endometriosis.

In this study, we aimed to investigate the genetics of heterogeneity in endometriosis by defining data-driven subtypes in women from the non-genotyped PMBB endometriosis population (N=4,078). We extracted clinical features known to be associated with endometriosis and performed unsupervised spectral clustering, identifying five clusters. Unsupervised clustering was an ideal approach for this study because it a way to find patterns in the data without introducing prior knowledge or bias. We chose spectral clustering with five clusters based on empirical metrics measured by comparing four different unsupervised clustering methods across a range of K values (2-20 clusters). This method had the best tradeoff between squared error, silhouette score, and cluster evenness.

To understand the clinical presentation patterns of each of the clusters, we compared the rates of the input features, diagnoses, and chart-reviewed phenotypes amongst them. Based on statistical enrichment testing across the features, the clusters were labeled as (1) pain comorbidities, (2) uterine disorders, (3) pregnancy complications, (4) cardiometabolic comorbidities, and (5) EHR-asymptomatic. This nuanced phenotyping, which diverges from traditional classifications, allows for a deeper understanding of the pathophysiological variations within endometriosis and highlights the necessity of tailored therapeutic approaches.

After deriving and characterizing the clusters in the non-genotyped PMBB, we used a k-nearest neighbors’ model to transfer the subtypes to the other four EHR-linked genetic datasets, PMBB, eMERGE, AOU, and UKBB. We performed ancestry-stratified candidate gene testing for each of the clusters using SAIGE and identified eight genome-wide significant signals. The genetic analysis of these clusters yielded intriguing results. While 46% of previously known GWAS loci were replicated in our study, significant differences in loci associations across the clusters were observed. For instance, genes like *WNT4* and *GREB1* showed specific associations with the uterine disorders and EHR asymptomatic clusters, suggesting that these genes might play distinct roles in the pathogenesis of these phenotypic presentations of endometriosis. Conversely, the *BSN* gene, although not statistically significant, demonstrated greater significance in the pain and pregnancy complications clusters, indicating a possible link to neurovascular or inflammatory mechanisms that could exacerbate these conditions.

Renalase (RNLS) is the protein associated with our only genome-wide significant association from the positive control, *RNLS/10q23*.*31*. At the Bonferroni significance threshold, the association with RNLS was significant for three out of five sub-phenotypes: pregnancy complications, cardiometabolic comorbidities, and EHR-asymptomatic. It was the only significant association with the cardiometabolic cluster. *RNLS* is highly expressed in the heart and contributes to regulating blood pressure (43). In genetic association studies, *RNLS* has been previously associated with type 1 diabetes (44) and smoking initiation (45). Smoking is a known risk factor of endometriosis.

Cluster three, with high rates of pregnancy-related complications such as infertility and high-risk pregnancy, was significantly associated with seven loci including *FSHB* (P = 1.8 x 10^-4^). The *FSHB* gene codes for the beta-subunit of follicle-stimulating hormone (FSH). FSH is essential for female fertility and has been shown to regulate myometrial contractile activity (46). *FSHB* was significantly associated with the positive control as well, but not with any of the other clusters.

Cluster five, which was largely asymptomatic in the EHR, was the largest cluster. Over half (56%) of UKBB endometriosis patients were assigned to this cluster. It is possible that those assigned to this cluster from any dataset have symptoms that were not recorded in the structured data which we had access to. Two well-known endometriosis loci from both of the last major GWASs are *SYNE1* and *CDKN2B-AS1* (16,17), both of which were significantly associated with the positive control and the EHR-asymptomatic cluster. Six loci were associated with this cluster and no other phenotypes: *BSN, ID4, CD109, MLLT10, IGF1*, and *SKAP1. MLLT10* and *BSN* have been previously associated with pain perception and maintenance (17). Serum levels of IGF-1 are significantly elevated in women with endometriosis (47). Gene expression of *ID4* is down-regulated in eutopic and ectopic endometrial tissue of women with endometriosis (48). *CD109I* and *SKAP1* have been previously associated with endometrial cancers (49,50).

Subtyping complex diseases, like endometriosis, is crucial for advancing precision medicine. The findings from our study underscore the utility of EHR as a rich resource for disease subtyping and genetic research. The linkage of detailed clinical data with genetic information enables the identification of phenotype-genotype correlations that are often diluted in broader GWAS analyses. Furthermore, the use of spectral clustering helps elucidate the heterogeneity within endometriosis, providing a framework for understanding the multifaceted nature of the disease and facilitating the development of personalized medicine.

However, it is essential to acknowledge the limitations of our study. One significant constraint was the sample size, which was particularly limited for some of the smaller clusters and for individuals of non-European ancestry. This limitation could potentially introduce bias and affect the generalizability of our findings. Additionally, our study relies on structured electronic health data only, which may not capture the full clinical picture and could be subject to inaccuracies or incomplete records. Lastly, this genetic association analyses in this study only focused on the candidate genes that are previously known to be associated with endometriosis. This approach might have restricted our ability to discover novel genetic loci potentially relevant to the specific clusters identified. Despite these limitations, our study marks a meaningful advancement in understanding the genetic factors that may contribute to the heterogeneity observed in endometriosis. By focusing on genetic associations gleaned from electronic health records, we offer a novel perspective that could be instrumental in future research and treatment approaches. To expand upon the current findings, future research should aim to perform comprehensive GWAS across the identified endometriosis subtypes. This will enable the detection of novel loci that could be crucial for understanding the distinct mechanisms underlying each subtype. Additionally, integrating multi-omics data (such as transcriptomic, proteomic, and metabolomic data) could further refine the molecular signatures associated with each cluster, enhancing the biological interpretability of the genetic associations. Another promising avenue is the longitudinal study of these clusters to assess disease progression and treatment outcomes, which could inform more effective, personalized therapeutic strategies.

In conclusion, our research highlights the importance of subtype-specific studies in elucidating the genetic basis of endometriosis. By leveraging the capabilities of EHR-linked biobanks and employing advanced clustering techniques, we pave the way for more targeted and effective approaches to understanding and managing this complex disease.

## Data Availability

The datasets generated during and/or analyzed during the current study are available from the corresponding author via collaborations upon request.

## Abbreviations

AOU: All of Us Biobank
eMERGE: electronic medical record and genomics network
EHR: electronic health record
GWAS: genome-wide association study
ICD: international classification of diseases
PMBB: Penn Medicine biobank
UKBB: United Kingdom biobank

## Acknowledgements

Research reported in this publication was supported by the Eunice Kennedy Shriver National Institute of Child Health and Human Development of the National Institutes of Health under award number R01HD110567.

We acknowledge the Penn Medicine BioBank (PMBB) for providing data and thank the patient-participants of Penn Medicine who consented to participate in this research program. We would also like to thank the Penn Medicine BioBank team and Regeneron Genetics Center for providing genetic variant data for analysis. The PMBB is approved under IRB protocol# 813913 and supported by Perelman School of Medicine at University of Pennsylvania, a gift from the Smilow family, and the National Center for Advancing Translational Sciences of the National Institutes of Health under CTSA award number UL1TR001878.

This phase of the eMERGE Network was initiated and funded by the NHGRI through the following grants: U01HG008657 (Group Health Cooperative/University of Washington); U01HG008685 (Brigham and Women’s Hospital); U01HG008672 (Vanderbilt University Medical Center); U01HG008666 (Cincinnati Children’s Hospital Medical Center); U01HG006379 (Mayo Clinic); U01HG008679 (Geisinger Clinic); U01HG008680 (Columbia University Health Sciences);

U01HG008684 (Children’s Hospital of Philadelphia); U01HG008673 (Northwestern University); U01HG008701 (Vanderbilt University Medical Center serving as the Coordinating Center); U01HG008676 (Partners Healthcare/Broad Institute); and U01HG008664 (Baylor College of Medicine).

We gratefully acknowledge All of Us participants for their contributions, without whom this research would not have been possible. We also thank the National Institutes of Health’s All of Us Research Program for making available the participant data examined in this study.

This research has been conducted using the UK Biobank Resource under Application Number 32133.

