## Supporting Information and Figures for "Enhancing Genetic Association Power in Endometriosis through Unsupervised Clustering of Clinical Subtypes Identified from Electronic Health Records"

### S1 Appendix: Clinical Features Extracted from the EHR

In total, we extracted 39 endometriosis-related phenotypes from the EHR. These 39 phenotypes included endometriosis anatomical subtypes, comorbidities, pregnancy-related features, and symptoms. Table S1 shows the prevalence of each feature and the ICD codes used to identify presence or absence. Some were used for clustering, and some were used for characterizing the clusters later.

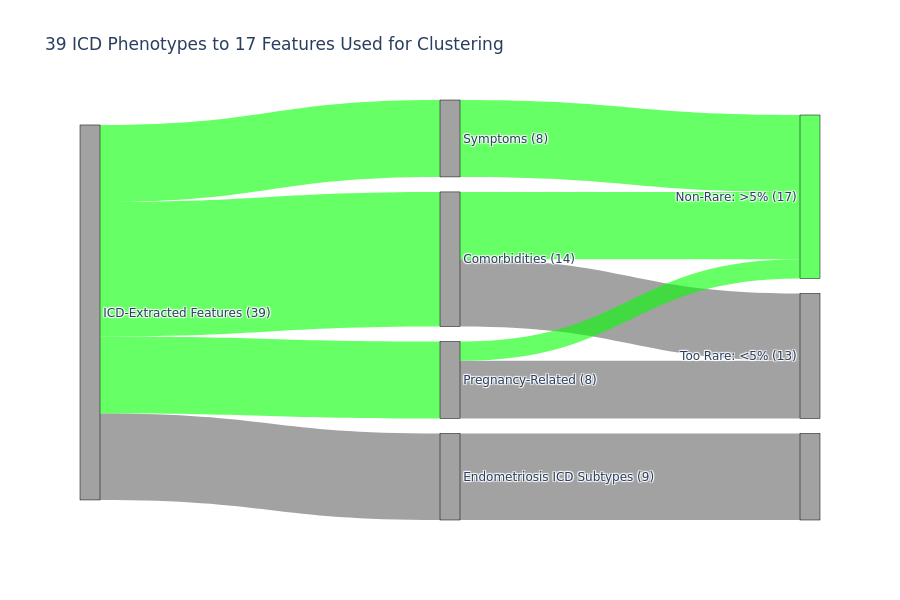

**Figure S1:** Sankey diagram showing the 39 endometriosis-related features used in our study. These were all extracted from the EHR using ICD-9 and ICD-10 codes. The green set on the top right with 17 features is the final set of features we used for clustering.

We selected the 17 clustering features based on their prevalence in the cluster training set, 4,078 endometriosis cases from the non-genotype PMBB. We extracted these 17 features for each of the genetic association test sets (PMBB, eMERGE, AOU, and UKBB) so that we could apply the same cluster classifier to them. Figure S2 shows the prevalence of each input feature across the five datasets.

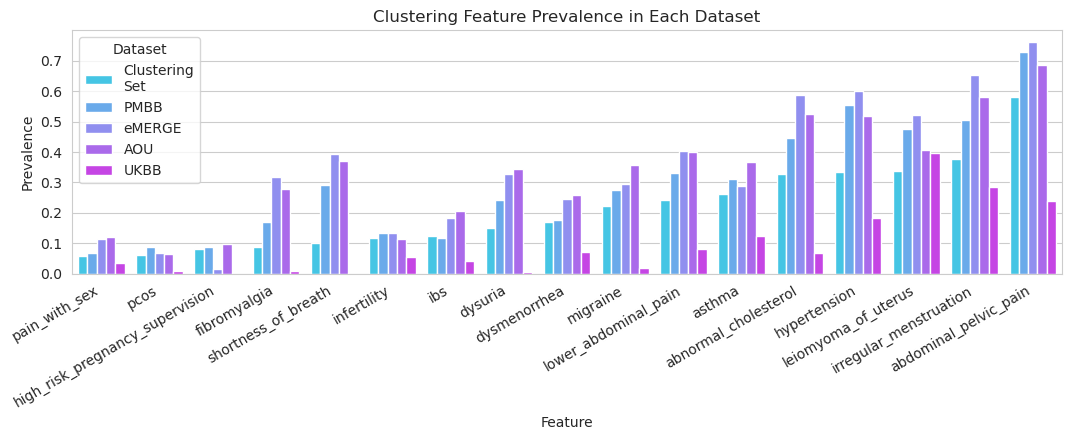

**Figure S2:** input feature prevalence from each dataset used in the study.

### S2 Appendix: Visual Methods Overview

A variety of datasets went into our study, and there were a few different roles that they served. The non-genotyped PMBB was used as the cluster derivation cohort. A subset of the genotyped PMBB underwent chart reviews to help characterize the clusters. The EHR-linked genetic datasets (Genotyped PMBB, eMERGE, AOU, and UKBB) were all used to test the genetic associations of the data-driven clusters. Figure S3 illustrates the process used for our study and how each dataset was incorporated.

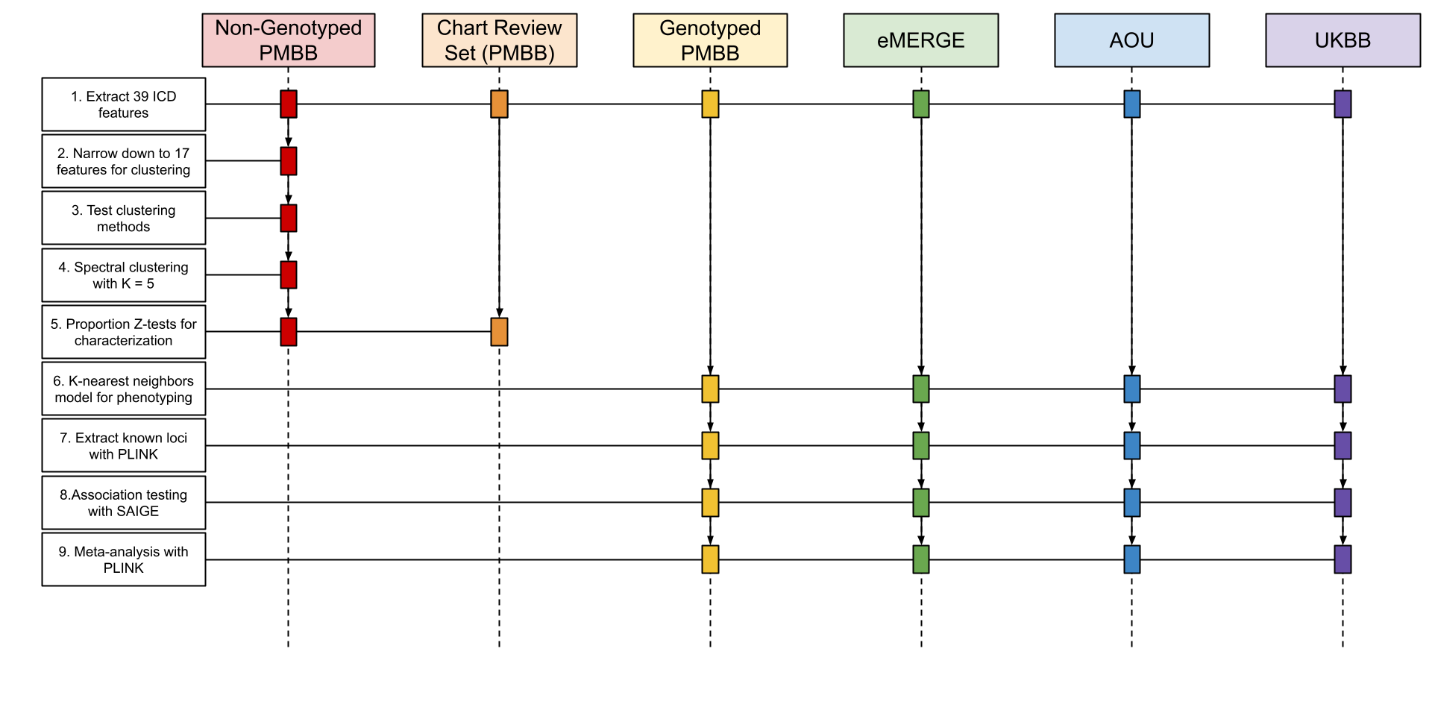

**Figure S3:** overview of the methods and how each dataset was utilized.
